# Molecular Profiling of COVID-19 Autopsies Uncovers Novel Disease Mechanisms

**DOI:** 10.1101/2021.04.04.21253205

**Authors:** Elisabet Pujadas, Michael Beaumont, Hardik Shah, Nadine Schrode, Nancy Francoeur, Sanjana Shroff, Clare Bryce, Zachary Grimes, Jill Gregory, Ryan Donnelly, Mary E. Fowkes, Kristin Beaumont, Robert Sebra, Carlos Cordon-Cardo

## Abstract

**Background:** Current understanding of COVID-19 pathophysiology is limited by disease heterogeneity, complexity, and a paucity of studies evaluating patient tissues with advanced molecular tools.

**Methods:** Autopsy tissues from two COVID-19 patients, one of whom died after a month-long hospitalization with multi-organ involvement while the other died after a few days of respiratory symptoms, were evaluated using multi-scale RNASeq methods (bulk, single-nuclei, and spatial RNASeq next-generation sequencing) to provide unprecedented molecular resolution of COVID-19 induced damage.

**Findings:** Comparison of infected/uninfected tissues revealed four major regulatory pathways. Effectors within these pathways could constitute novel therapeutic targets, including the complement receptor C3AR1, calcitonin-like receptor or decorin. Single-nuclei RNA sequencing of olfactory bulb and prefrontal cortex highlighted remarkable diversity of coronavirus receptors. Angiotensin I converting enzyme 2 was rarely expressed, while Basignin showed diffuse expression, and alanyl aminopeptidase was associated with vascular/mesenchymal cell types. Comparison of lung and lymph node tissues from patients with different symptomatology with Digital Spatial Profiling resulted in distinct molecular phenotypes.

**Interpretation:** COVID-19 is a far more complex and heterogeneous disease than initially anticipated. Evaluation of COVID-19 rapid autopsy tissues with advanced molecular techniques can identify pathways and effectors at play in individual patients, measure the staggering diversity of receptors in specific brain areas and other well-defined tissue compartments at the single-cell level, and help dissect differences driving diverging clinical courses among patients. Extension of this approach to larger datasets will substantially advance the understanding of the mechanisms behind COVID-19 pathophysiology.

**Funding:** No external funding was used in this study.

**Research in context:** *Evidence before this study:* Information regarding changes seen in COVID-19 has accumulated very rapidly over a short period of time. Studies often rely on examination of normal samples and model systems, or are limited to peripheral blood or small biopsies when dealing with tissues collected from patients infected with SARS-CoV-2. For that reason, autopsy studies have become an important source of insights into the pathophysiology of severe COVID-19 disease, highlighting the emerging role of hyperinflammatory and hypercoagulable syndromes. Studies of autopsy tissues, however, are usually limited to histopathologic and immunohistochemical evaluation. The next frontier in understanding COVID-19 mechanisms of disease will require generation of highly dimensional, patient-specific datasets that can help dissect this complex and heterogeneous disease.

*Added value of this study:* Our work illustrates how high-resolution molecular and spatial profiling of COVID-19 patient tissues collected during rapid autopsies can serve as a hypothesis-generating tool to identify key mediators driving the pathophysiology of COVID-19 for diagnostic and therapeutic target testing. Here we employ bulk RNA sequencing to identify key regulators of COVID-19 and list specific mediators for further study as potential diagnostic and therapeutic targets. We use single-nuclei RNA sequencing to highlight the diversity and heterogeneity of coronavirus receptors within the brain, suggesting that it will be critical to expand the focus from ACE2 to include other receptors, such as BSG and ANPEP, and we perform digital spatial profiling of lung and lymph node tissue to compare two patients with different clinical courses and symptomatology.

*Implications of all the available evidence:* COVID-19 is a far more heterogeneous and complex disease than initially anticipated. Advanced molecular tools can help identify specific pathways and effectors driving the pathophysiology of COVID-19 and lead to novel biomarkers and therapeutic targets in a patient-specific manner. Larger studies representing the diversity of clinical presentations and pre-existing conditions will be needed to capture the full complexity of this disease.

## Introduction

Concerted research efforts are quickly advancing our understanding of COVID-19 pathophysiology, highlighting pathways that influence uncontrolled cytokine release and vascular injury^1,2,3^, and suggesting distinct disease stages^4^. Studies directly evaluating patient tissues^5,6,7^, however, often lack the resolution afforded by interrogation with advanced molecular methods. Highly dimensional, patient-specific datasets can be generated through multi-scale, next-generation sequencing of well-preserved rapid autopsy tissues. Bulk sequencing yields high-read depth profiling, while single-nuclei methods and spatial profiling can interrogate distinct cellular subpopulations. Mining of such datasets can facilitate novel hypothesis generation, thus advancing the molecular understanding of SARS-CoV-2 infection and COVID-19 disease mechanisms.

## Methods

### Tissue collection

Rapid autopsies (6 hours) were performed with appropriate consent and following COVID-19 safety protocols^8^. Fresh tissues were frozen on dry ice and stored at −80°C. Formalin-fixed paraffin-embedded tissues (FFPE) were collected and processed following routine protocols.

### Bulk RNASeq and Gene Set Enrichment Analysis (GSEA)

Bulk RNASeq was performed from two technical replicates across ten tissues harvested from Patient 1 (see Supplemental Methods).

### Single-nuclei RNASeq (snRNASeq)

Debris-free nuclei (∼10,000) were isolated from frozen olfactory bulb (OB) and prefrontal cortex (PFC) tissue from Patient 1. Paired-end cDNA libraries were prepared using the 10x Chromium platform (v3 protocol) and sequenced on a NovaSeq instrument (Illumina) to a read depth of 50,000-100,000 reads per nucleus. Cell types were annotated through machine learning based on published data^9^ (see Supplemental Methods).

### Digital Spatial Profiling (DSP)

Unstained 4µm-thick sections were cut from FFPE blocks from two patients and immunoflourescently labeled with PanCK, CD45, CD68 and DAPI. Twelve regions of interest (ROIs) per slide were selected for targeted spatial transcriptomics (GeoMx, Nanostring) with >1,600 cancer atlas transcripts and 10 additional SARS-CoV-2 genes (see Supplemental Methods).

## Results

Fresh-frozen tissue from several organs was obtained at the time of autopsy for Patient 1, a male in his 60s with a complex medical history including HIV, status post-transplant and cancer in clinical remission whose COVID-19 hospital admission lasted over a month (**Figure 1A**). Bulk RNASeq evaluation revealed viral RNA in the nasopharynx and lung (148 and 120 reads, respectively, mapping to SARS-CoV-2 reference genome), but not in OB, PFC, oropharynx, salivary gland, tongue, heart, liver or kidney (**Figure 1B**). Differential gene expression analysis between infected and uninfected tissues resulted in 4,376 differentially expressed genes (FDR <0.05). To assess biological significance, GSEA was performed to probe for Gene Ontology (GO) categories significantly overrepresented in infected versus uninfected tissues (FDR < 0.05) (**Figure 1C**). The top four connected GO categories include “blood vessel development”, “cytokine production”, “cell activation”, and “structure and degradation”. All four pathways share TNFα induced protein 3 (TNFAIP3), complement 3a anaphylatoxin chemotactic receptor (C3AR1), and ADAM8, a metalloproteinase involved in leukocyte extravasation. Other examples of conserved molecules across multiple pathways include angiopoietin 1 (ANGPT1), calcitonin-like receptor (CALCRL), and decorin (DCN), an activator of autophagy.

**Figure 1 –.**
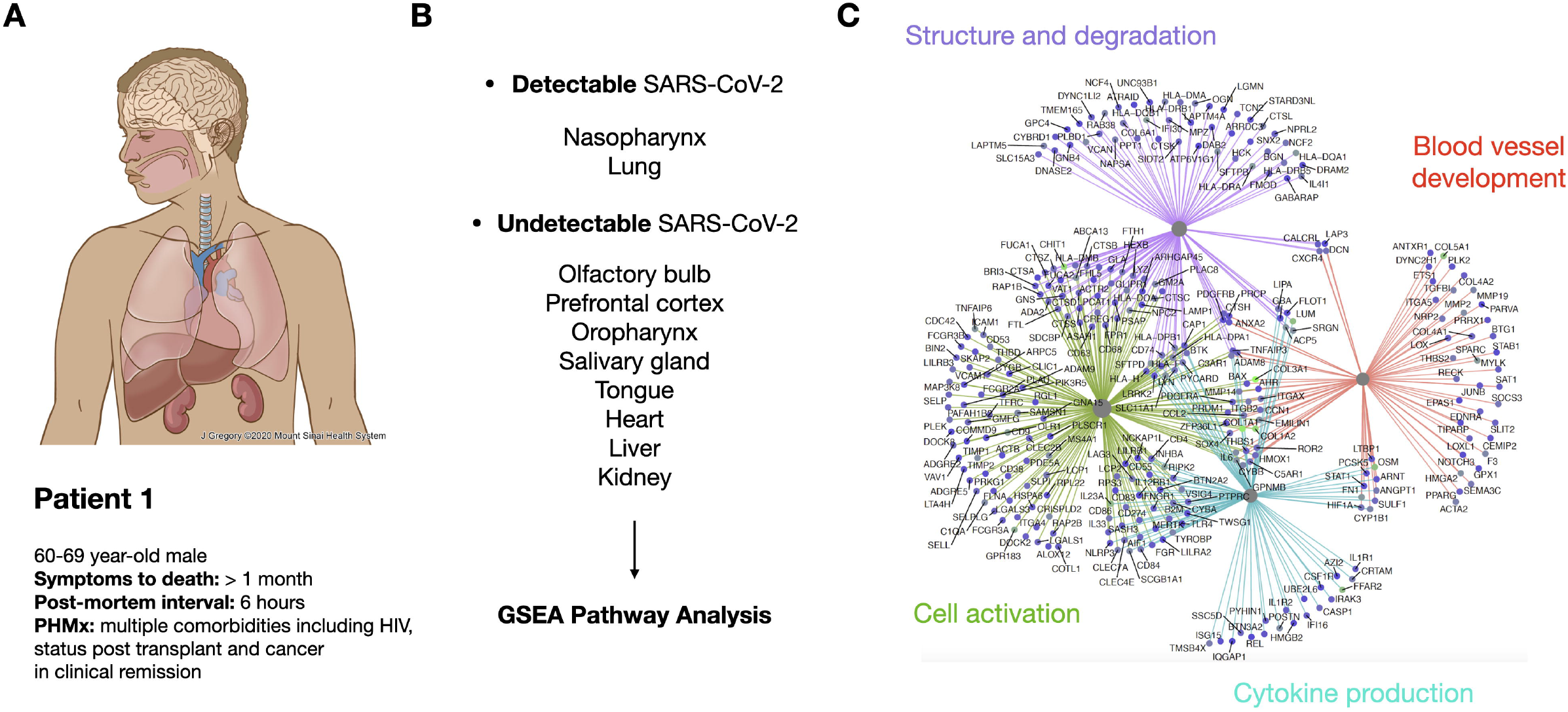
Bulk RNASeq and GSEA Pathway Analysis. **(A)** Samples from multiple organs were collected at the time of autopsy for Patient 1, a male in his 60s with a complex medical history and a long disease course. **(B)** Reads mapping to the SARS-CoV-2 genome in unenriched host tissue were detected in nasopharynx and lung, while no reads were detected in olfactory bulb, pre-frontal cortex, oropharynx, salivary gland, tongue, heart, liver or kidney. Organ-specific datasets were group by infected-uninfected status for further analysis. **(C)** Visualization of Gene Set Enrichment Analysis (GSEA) showing the top 4 most statistically significant pathways. Nodes represent specific transcripts and edges are color-coded to reflect the pathways where the node is enriched. Nodes can have edges from multiple pathways.

snRNASeq was performed on OB and PFC of Patient 1 (**Figure 2A**). Dimensionality reduction and clustering showed expected separation between OB and PFC (**Figure 2B**) and identified 25 unique subpopulations (**Figure 2C**) that were annotated using existing databases (**Figure 2D**, see Supplemental Methods). Expression levels of any transcript from nuclei within these subpopulations can inform hypotheses unresolvable by bulk RNA analyses. For example, examination of multiple coronavirus-associated receptors reveals only scattered expression of angiotensin I converting enzyme 2 (ACE2) in rare cells and robust expression of Basignin (BSG) throughout, while alanyl aminopeptidase (ANPEP) is exclusively expressed in two subpopulations enriched for pericytes, vascular, fibroblastic, and stromal cells (**Figure 2E)**.

**Figure 2 –.**
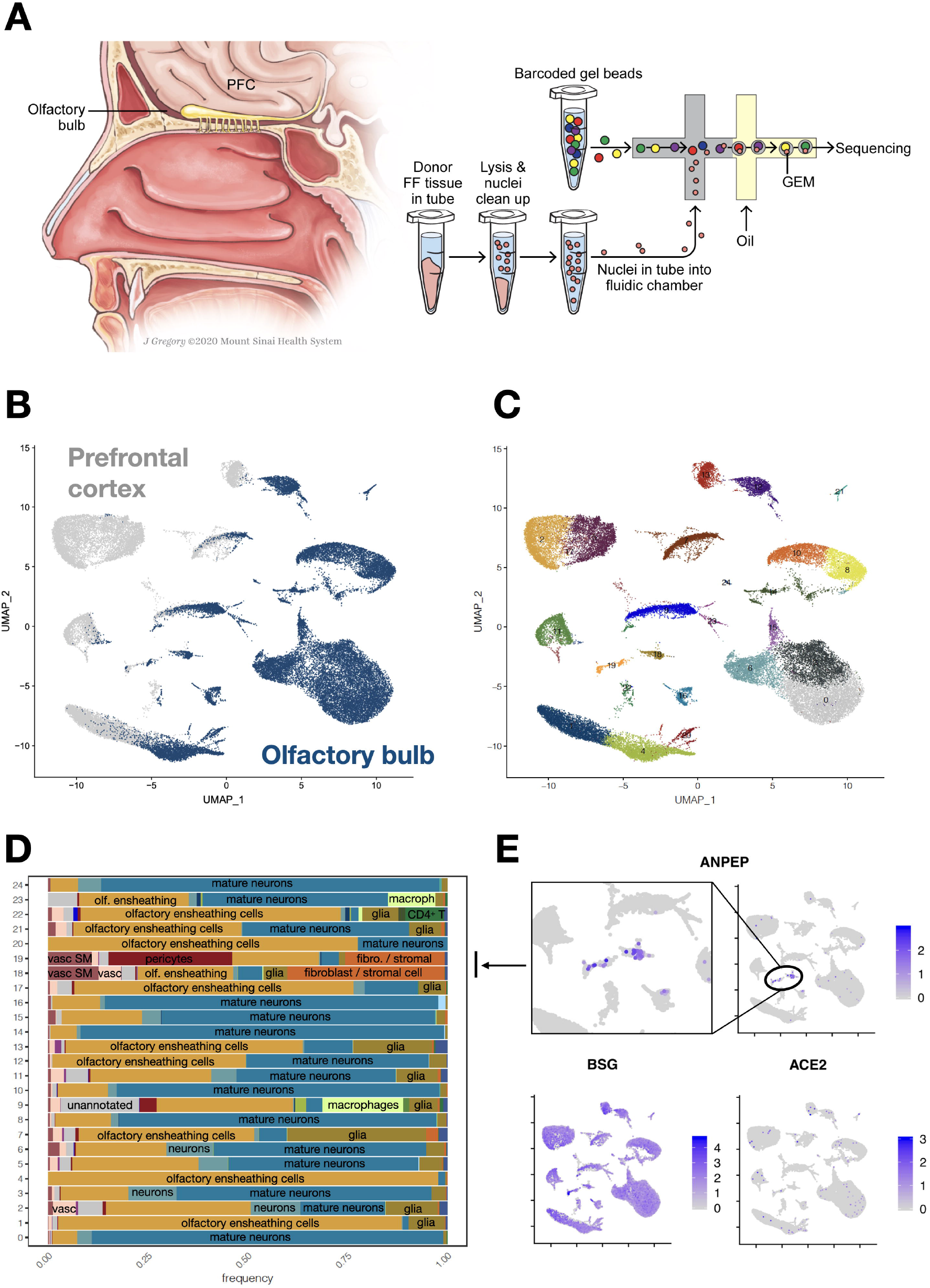
Single-nuclei sequencing of olfactory bulb and prefrontal cortex. **(A)** Anatomical illustration indicating the location of the olfactory bulb and the overlying pre-frontal cortex (PFC) collected from Patient 1 (left). Diagram demonstrating 10x Genomics technology for single-nuclei sequencing. Briefly, fresh-frozen (FF) tissue is lysed, nuclei are cleaned up and flowed into a microfluidic chamber where they combine with barcoded gel beads to generate GEMs (Gel-Bead in Emulsions) that are subsequently amplified and sequenced (right). **(B)** Uniform Manifold Approximation and Projection (UMAP) dimensionality reduction graph shows nuclei subpopulations where prefrontal cortex origin is labeled in gray and olfactory bulb origin is labeled in navy blue, showing anatomically distinct subcompartments as well as clusters with a mixture of cell populations. **(C)** Uniform Manifold Approximation and Projection (UMAP) dimensionality reduction graph illustrating 25 distinct nuclei subpopulations based on their transcriptional profiles. **(D)** Cell types for each nuclei subpopulation were annotated used machine learning (see Supplemental Methods). The x-axis shows the frequency of each annotated cell type, while the y-axis has each distinct nuclei subpopulation shown in B and C. As expected, given the anatomical source of these tissues, a majority of cells types fall under the categories of olfactory ensheathing cells, neurons and mature neurons. There are also less frequent but important cell types such as glia, macrophages, CD4^+^ T lymphocytes, fibroblast and stromal cells, vascular cells, vascular smooth muscle cells and pericytes. **(E)** Exploration of the expression of coronavirus-related receptors ACE2 (bottom right), BSG (bottom left) and ANPEP (top), with a higher-power view of ANPEP in clusters 18 and 19 (top left). The arrow connects the two highlighted clusters (18 and 19) with increased expression of ANPEP with their annotation in panel D.

Digital Spatial Profiling (DSP) was performed on FFPE tissue across multiple organs from two COVID-19 autopsy patients. Patient 1, as above, and Patient 2, a male in his 60s with diabetes and heart failure who died after only 3 days of symptoms (**Figure 3A**). Following immunostaining, 12 ROIs per slide (**Figures 3B,D**) were selected for targeted spatial sequencing. Viral transcripts (ORF1ab and S) were detected in the lungs of Patient 2 (**Figure 3C**), along with a more robust inflammatory response compared to Patient 1, including higher expression of MX1, an interferon-induced GTPase with antiviral activity against RNA viruses, interleukin-1 receptor-like 1 (IL1IR1), and interferon alpha-inducible protein 6 (IFI6), a negative regulator of apoptosis (**Figure 3C**). Comparison of lymph node ROIs between these two patients showed diverging inflammatory signatures (**Figure 3E**). Patient 1 had higher expression of the chemotactic factors CCL15 and CCL18, along with TNF and ITGB2, a receptor for the iC3b complement fragment and fibrinogen, while Patient 2 had higher expression of transcripts related to cell proliferation and division, including thymidylate synthetase (TYMS), CCNA2, NASP, lymphocyte antigen 6E (LY6E) and multiple histone subunits (**Figure 3E**).

**Figure 3 –.**
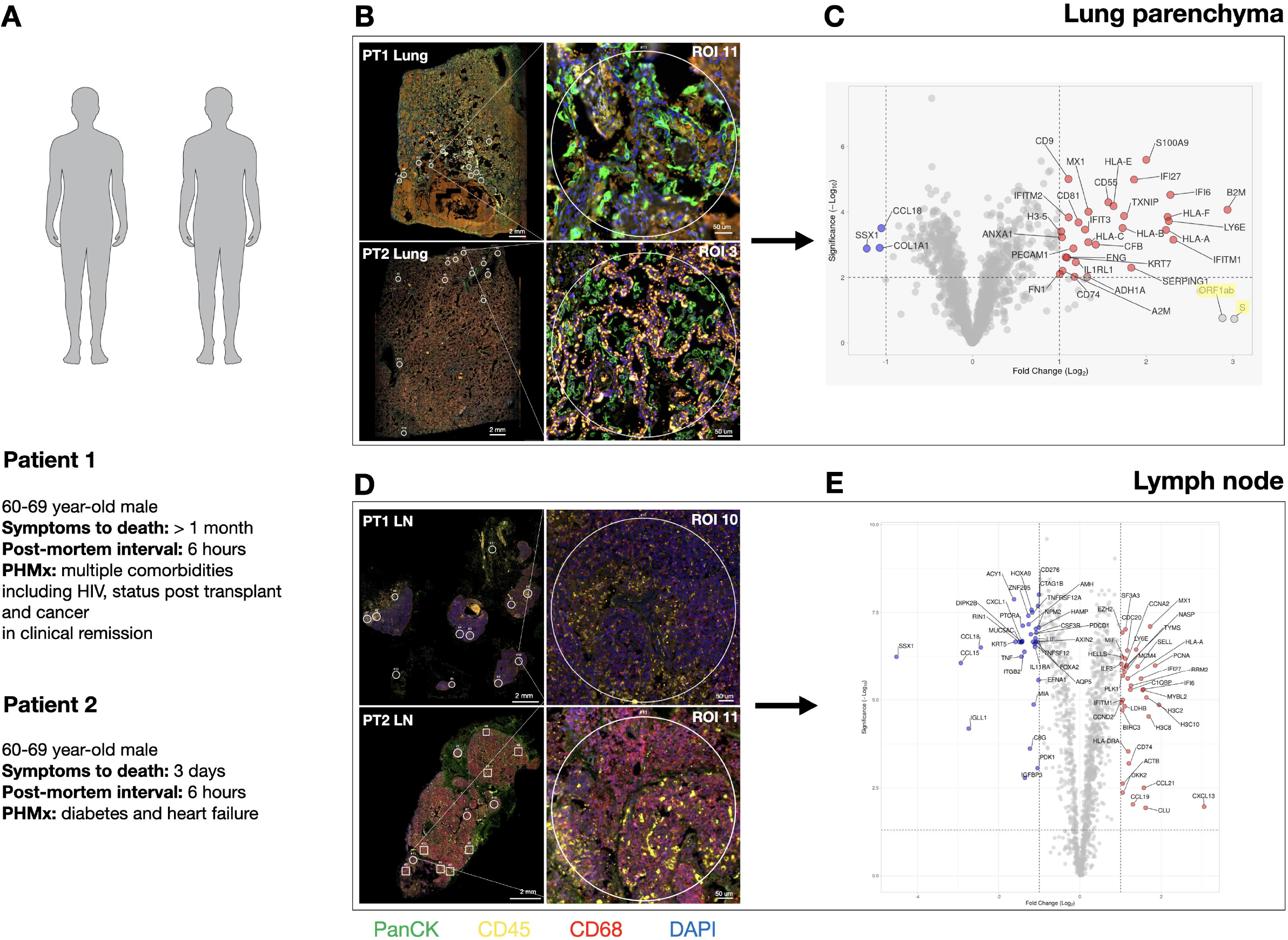
Digital Spatial Profiling of lung and lymph nodes from Patients 1 and 2. **(A)** Summary of relevant clinical details for Patient 1 and Patient 2. **(B)** Whole slide images of immunostained FFPE lung sections from Patient 1 (top left) and Patient 2 (bottom left). Small circles denote selected ROIs. Representative ROIs are highlighted: ROI 11 from Patient 1 (top right) and ROI 3 from Patient 2 (bottom right). **(C)** Volcano plot comparing gene expression profiles using the commercial Nanostring GeoMx platform. Transcripts labeled in blue are significantly more expressed in Patient 1, while transcripts labeled in red are significantly more expressed in Patient 2. Yellow highlights denote viral transcripts. **(D)** Whole slide images of immunostained FFPE lymph node sections from Patient 1 (top left) and Patient 2 (bottom left). Small circles denote selected ROIs. Representative ROIs are highlighted: ROI 10 from Patient 1 (top right) and ROI 11 from Patient 2 (bottom right). **(E)** Volcano plot comparing gene expression profiles using the commercial Nanostring GeoMx platform. Transcripts labeled in blue are significantly more expressed in Patient 1, while transcripts labeled in red are significantly more expressed in Patient 2.

## Discussion

Here we present multi-scale, high resolution molecular profiling of select tissues from COVID-19 patients collected at autopsy and demonstrate how these technologies can facilitate hypothesis generation. First, we employed bulk RNASeq to evaluate presence of viral RNA across organs from Patient 1; then compared infected and uninfected tissues to determine key regulatory pathways associated with infection, including “blood vessel development”, “cytokine production”, “cell activation”, and “structure and degradation”. Numerous specific regulators are listed including C3AR1, a complement receptor controlling neutrophil extracellular trap (NET) formation that can lead to a pro-thrombotic state^10^, both important mechanisms implicated in COVID-19 progression^4^. C3AR1 is thus one of many examples that could serve as valuable target.

We next performed snRNASeq in the OB and PFC of Patient 1. While significant attention has been directed to ACE2, there are numerous coronavirus-associated receptors^11^. In the brain tissues analyzed here we found that ACE2 was rarely expressed, while BSG was more widely expressed, and ANPEP showed preferential expression in vascular cell types, such as endothelial cells and pericytes. Understanding the true complexity of SARS-CoV-2 infection and COVID-19 disease will require careful mapping of all receptors and factors in a manner that can resolve each cellular niche.

Lastly, we demonstrate DSP on FFPE tissues, emphasizing the importance of integrating transcriptional profiling with histologic features and microanatomical detail. This technology allows evaluation of a larger sample set, yet still provides viral detection and the ability to compare patients with different characteristics and disease courses. While many factors could account for the differences seen between our two patients, it is tempting to speculate that some of our findings reflect early-versus late-stage COVID-19, both in terms of viral presence and robustness of lymphocytic activation in Patient 2, with a shorter disease course; and phagocytic and coagulopathic signatures in Patient 1, with a prolonged hospitalization and extended organ damage. The limited cohort size and the complexity of the clinical histories precludes further generalization of these findings.

In summary, we describe four key pathways in severe COVID-19 and provide specific effectors for further study as potential diagnostic and therapeutic targets. In addition, we reveal remarkable heterogeneity of coronavirus receptors within well-defined anatomic compartments, and diverging molecular signatures between patients with different clinical courses, underscoring the need to deploy advanced molecular techniques to dissect the complexity of COVID-19 pathophysiology.

## Supporting information

IRB exemption letter

## Data Availability

Datasets generated and analyzed in this study will be made available through Gene Expression Omnibus (GEO).

## Contributors

EP: conceptualization, resources, data curation, methodology, investigation, writing original draft, writing – review & editing. MB: conceptualization, data curation, formal analysis, visualization, investigation, writing – review & editing. HS,NS,NF,SS: data curation, formal analysis, visualization, investigation, writing – review & editing. CB,ZG,RD: resources, methodology, investigation, data curation. JG: data curation, visualization. MF: conceptualization, resources, methodology, investigation, data curation. KB: conceptualization, data curation, formal analysis, visualization, investigation, writing – review & editing. RS: conceptualization, resources, data curation, formal analysis, visualization, investigation, methodology, supervision, writing original draft, writing – review & editing. CCC: conceptualization, investigation, resources, data curation, visualization, methodology, supervision, writing original draft, writing – review & editing.

