## Supplemental Methods for "Molecular Profiling of COVID-19 Autopsies Uncovers Novel Disease Mechanisms"

**Uncovers Novel Disease Mechanisms**

**Supplemental Methods:**

**Bulk RNA sequencing and Gene Set Enrichment Analysis (GSEA).** Illumina RNA sequencing was performed on two technical replicates across ten tissues harvested from a single rapid autopsy (Patient 1). The raw RNAseq data was aligned to the hg38 genome using the *STAR* mapper, annotated using GENCODE v35 gene annotations and counted using *featureCounts*. Genes with fewer than 50 counts across all samples were excluded from the analysis, and the counts were subsequently log2-transformed for differential gene expression analysis using *DESeq2.* The SARS-CoV-2 reference genome employed here can be found in GISAID under accession EPI_ISL_414476. A pairwise comparison between infected tissues (lung, nasopharynx) and uninfected tissues (olfactory bulb, prefrontal cortex, oropharynx, salivary gland, tongue, heart, liver, kidney) resulted in 4,376 differentially expressed (DE) genes with an FDR < 0.05. To assess the biological significance of the DE genes, a Gene Set Enrichment Analysis (*clusterProfiler*) was performed to probe for Gene Ontology (GO) categories that were significantly overrepresented and statistically different in infected versus non-infected tissues. The top four (FDR < 0.05) connected GO categories were plotted as a network linking genes to biological functions and revealed cell activation (green), lytic vacuole (purple), cytokine production (blue) and blood vessel development (red) as overrepresented pathways. Large gray nodes represent GO term biological processes, smaller nodes represent DE genes in infected versus uninfected tissues and are shaded according to fold change size.

**Single-nuclei isolation and snRNAseq of OB and PFC tissues.** Nuclei were isolated from frozen tissue following the recommended protocol from 10x Genomics. In short, lysis buffer (10 mM Tris-Hcl, 10 mM NaCl, 3 mM MgCl2 and 0.1% Nonident P40 substitute in nuclease free water) was added to frozen tissue, which was gently homogenized. Following centrifugation, nuclei were washed in PBS + 0.04% BSA and strained to remove aggregates. Single nuclei RNA Seq was performed on these samples using the Chromium platform (10x Genomics, Pleasanton, CA) with the 3’ gene expression (3’ GEX) V3 kit, and an input of ~10,000 nuclei from a debris-free suspension. Briefly, Gel-Bead in Emulsions (GEMs) were generated on the sample chip in the Chromium controller. Barcoded cDNA was extracted from the GEMs by Post-GEM RT-cleanup and amplified for 12 cycles followed by fragmentation of amplified cDNA, end-repair, poly A-tailing, adapter ligation, and 10X-specific sample indexing following the manufacturer’s protocol. Libraries were quantified using QuBit (Thermofisher) and Bioanalyzer (Agilent). Libraries were sequenced in paired end mode on a NovaSeq instrument (Illumina, San Diego, CA) targeting a depth of 50,000-100,000 reads per nucleus.

The Cell Ranger Single-Cell Software Suite (version 3.1, 10x Genomics) was used to align and quantify sequencing data against the provided GRCh38 human reference genome. Downstream analyses, such as graph-based clustering, and differential expression analysis/visualization, was performed using the Loupe Cell Browser (version 3.0, 10x Genomics). Then, count matrices of all samples were imported and prepared using the R Seurat package^1^. Quality control for doublets and low-quality cells was achieved through exclusion of cells with less than 200 or more than 5000 transcripts and those with a higher than 20% mitochondrial gene contribution, respectively. Count data was log-normalized and transcripts were scaled and centered, using built-in Seurat functions. Then, filtered and normalized counts of all samples were merged using the Seurat-specific merge() function.

Next, dimensionality reduction and clustering were performed where variable transcripts were calculated based on standardized feature values using observed mean and expected variance of a local polynomial regression model. On the resulting variable transcripts 20 principal components were computed, which in turn were used as input for uniform manifold approximation and projection (UMAP) dimensionality reduction. For clustering analysis, a shared nearest neighbor (SNN) graph was performed, and the modularity function optimized using the Leiden algorithm. Last, cell types were identified and annotated automatically through machine learning using the singleCellNet R package^2^, as described in the developer’s manual. Briefly, a Random Forest classifier was trained on a scRNA-seq data set of olfactory neuroepithelium from a healthy donor, following cell type annotation^3^ and assessed on a withheld subset thereof. The classifier was then applied to the current data set and a cell type was predicted for each cell.

**Digital Spatial Profiling (DSP) data generation and annotation:** Unstained sections were freshly cut from FFPE blocks at 4µm-thickness onto positively charged slides, stored at 4ºC and immunoflourecent tissue profiling was performed using PanCK (488) cytokeratin, CD45 (647) immune response, CD68 (568) macrophage inflammatory response, and DAPI DNA staining to enable selection of 12 regions of interest (ROIs) for targeted spatial transcriptomics using the commercial Nanostring GeoMx. Circular ROIs measured 600µm in diameter and square ROIs measured 600x600 µm. The spatial transcriptomic panel included >1600 targeted cancer transcriptome atlas panel with 10 additional custom SARS-CoV-2 genes added (Ace2-mM, ACE2-Hs, E, M, N, orf1a, ORF3a, ORF7a, ORF8 and S protein) to encompass a comprehensive detection of host immune and inflammatory response alongside direct SARS-CoV-2 infection genes. Once DSP data quality control was completed, downstream differential expression of 6 ROIs (3 high and 3 low inflammation) from each patient were concatenated and differentially expressed genes between patients were determined to demonstrate the ability to differentiate hallmark infection, inflammatory and/or immune response between the two patients in this study. Briefly, reads were processed for high quality with adapter removal resulting in trimmed reads, and the paired-end reads were merged and aligned. Then PCR duplicates were removed by matching on the Unique Molecular Index (UMI), resulting in deduplicated reads. Then, QC was done in two steps, first Segment QC and then Biological Probe QC.  In segment QC, raw read threshold, percent aligned reads, sequence saturation and technical background were compared to an expected range to determine the overall quality of the data. We then performed Biological Probe QC to determine the thresholds for excluding outlier probes. Once all ROIs were processed, the raw counts were Q3 normalized against housekeeping genes by averaging to the geometric mean of the internal spike-in controls by which system variation was accounted for. Desired ROIs were annotated with their inflammatory and pathologic status and grouped respectively. The volcano plot illustration shows results of a paired t-test of ROIs from each patient tissue section.
