## Supplementary material for "Molecular Profiling of COVID-19 Autopsies Uncovers Novel Disease Mechanisms": IRB exemption letter

Mount Sinai

Icahn School of Medicine at Mount Sinai
Mount Sinai Beth Israel
Mount Sinai Brooklyn
The Mount Sinai Hospital
Mount Sinai Queens
New York Eye and Ear Infirmary
of Mount Sinai
Mount Sinai St. Luke's
Mount Sinai West

**Program for the Protection of Human Subjects**
*Institutional Review Boards*

Mount Sinai Health System
One Gustave L. Levy Place, Box 1081
New York, NY 10029-6574
T 212-824-8200
F 212-876-6789

icahn.mssm.edu/pphs

March 29, 2021

**Carlos Cordon-Cardo, MD, PhD**
Chairman, Pathology, Molecular & Cell Based Medicine
Annenberg Building
1468 Madison Ave, 15-60
New York, NY 10029

**RE: IRB Broad Position Statement Regarding Use of Autopsy Material for Research**

Dear Carlos,

I am writing to corroborate that the Icahn School of Medicine at Mount Sinai, through the Institutional Review Board (IRB) of the institution, considers autopsy materials as "not human subjects research" and do not fall under the purview of the IRB. The acquisition of high quality tissues at time of autopsy has been fundamental to support numerous institutional research efforts to understand the pathophysiology of human disease, most recently in the context of the COVID-19. Facilitating this research is a goal to which I have committed the Icahn School of Medicine at Mount Sinai's Program for the Protection of Human Subjects, and which I consider to be of exceptionally high value to the community at large.

Best regards,

Glenn Martin, MD, CIP
Senior Associate Dean for Human Subjects Research
Executive Director Program for the Protection of Human Subjects
Icahn School of Medicine at Mount Sinai
